# Descriptive Epidemiology of Kyasanur Forest Disease in Thirthahalli Taluk, Shivamogga, 2018-2022

**DOI:** 10.1101/2024.04.26.24306123

**Authors:** KV Srividya, LR Bhavesh, Sushma Choudhary, Ajit Shewale, DM Pallavi, MR Padma, Simmi Tiwari

**Author notes:** Corresponding author: Dr. Srividya KV, MBBS, MS, India EIS Fellow, EIS Cell, Epidemiology Department, National Centre for Disease Control, Sham Nath Marg, New Delhi 110054.

## Abstract

**Background:** Kyasanur Forest Disease (KFD) is a zoonotic viral disease caused by the Kyasanur Forest Disease Virus (KFDV), belongs to the family Flaviviridae, and is transmitted to humans through the bites of infected ticks, primarily in forested regions. Thirthahalli taluk from Shivamogga district of Karnataka has been reporting consistently high burden of KFD. We describe the epidemiology of KFD cases in Thirthahalli taluk including their temporal and spatial distribution, correlate KFD hot-spots with the number of cases and identify high-risk groups.

**Methods:** We conducted a retrospective analysis of KFD surveillance data obtained from district surveillance unit, Shivamogga, on human, tick and monkey data, of Thirthahalli taluk from November 2017 to May 2022, described epidemiology, calculated the annual and seasonal time-trend of cases and correlated number of cases with number of tick pools and/ or monkey carcass positive for KFDV. We report Pearson’s correlation coefficient. We performed spatial analysis to identify clustering patterns and describe the PHC-wise disease burden.

**Results:** Thirthahalli taluk reported 326 cases, all were lab-confirmed using either RT-PCR (93%) or IgM-ELISA (7%); with an annual incidence rate of 45.9 per 100,000 population, 58% in males and median age 40 (IQR 28-51) years. Most (71,5%) cases occurred from February to April, case fatality rate was 1.8%. The maximum number of cases occurred in the years 2019 (38%) and 2020 (44%) crossing the outbreak threshold. The populations coming under six Primary Health Centre (PHCs) reported 84% of all cases and reported all deaths. There was correlation between number of human cases in a PHC and tick pool positivity (r=0.62); and positive tick pools and positive monkey carcasses (r=0.71). Age groups with the highest average annual incidence were 45 to 49 years (8.4 per 100,000 population) and 50-54 years (8.1 per 100,000 population).

**Conclusion:** Thirthahalli taluk had an ongoing endemic transmission of KFD during the study period including two outbreaks in consecutive years. All deaths occurred in PHCs with a high case burden; occurrence of human cases per year was moderately correlated with tick pool positivity. Age groups most at risk for both the occurrence of the disease and mortality were middle-aged adult males. We recommend continued tick surveillance and monkey carcass testing to map disease hot-spots; and targeted interventions in high-burden PHCs and at-risk age groups for effective disease control.

## Introduction

Kyasanur Forest Disease (KFD) is a tick-borne viral haemorrhagic fever, first reported from Shivamogga district, Karnataka state, in 1957 as an outbreak of acute febrile illness accompanied with altered sensorium, seizures and haemorrhage among the people residing in ‘Kyasanur forest’ area of Shivamogga district. ^1,2^ The KFD virus (KFDV) belongs to the family Flaviviridae, and is spread via the bite of infected ticks belonging to the species *Haemaphysalis spinigera*.^3^ KFD causes epizootics in primates resulting in high fatality, with monkey death regarded as an early signal of KFDV transmission in that area.^4^ Areas reporting *Haemaphysalis* ticks and/ or monkey carcass infected with KFDV are considered disease hot-spots and prioritized for surveillance and control activities.^5^ The transmission season is from late November to June during the active period of nymphal *Haemaphysalis* ticks (figure 1).^6^ Since the first report in 1957, regular KFD outbreaks have been reported in Karnataka with 400-500 cases annually and a case fatality rate of 3-10%.^4,7^ In view of the public health threat from this emerging zoonotic disease, Karnataka state has instituted KFD surveillance in the affected areas to monitor, prevent and control KFD cases and outbreaks. Key strategies of the program include surveillance of human cases, monkey deaths and tick pool positivity along with KFD vaccination and vector control measures in cattle.^8^ The KFD vaccine is a formalin-inactivated vaccine, with two primary doses (one month apart) and a booster (after 6-9 months of the second primary dose), given to the eligible population aged six years and above. Annual boosters are recommended every year in the at-risk population. The program defines at-risk population as people living within a 5 Km aerial distance of a lab-confirmed KFD human positive, tick-pool positive or monkey death^8^. In spite of these measures, Shivamogga continues to record KFD cases and deaths every year. Thirthahalli taluk (figure 2) (sub-district), one of the seven taluks under Shivamogga district, has been reporting high case burden consistently.^9,10^ However, no studies on the descriptive epidemiology of KFD from Thirthahalli have been published in literature. Hence, in this study we describe the epidemiology of KFD cases in Thirthahalli taluk including their temporal and spatial distribution, correlate number of cases with the number of tick pools and/ or monkey carcass positive for KFDV, and identify high-risk groups.

**Figure 1.**
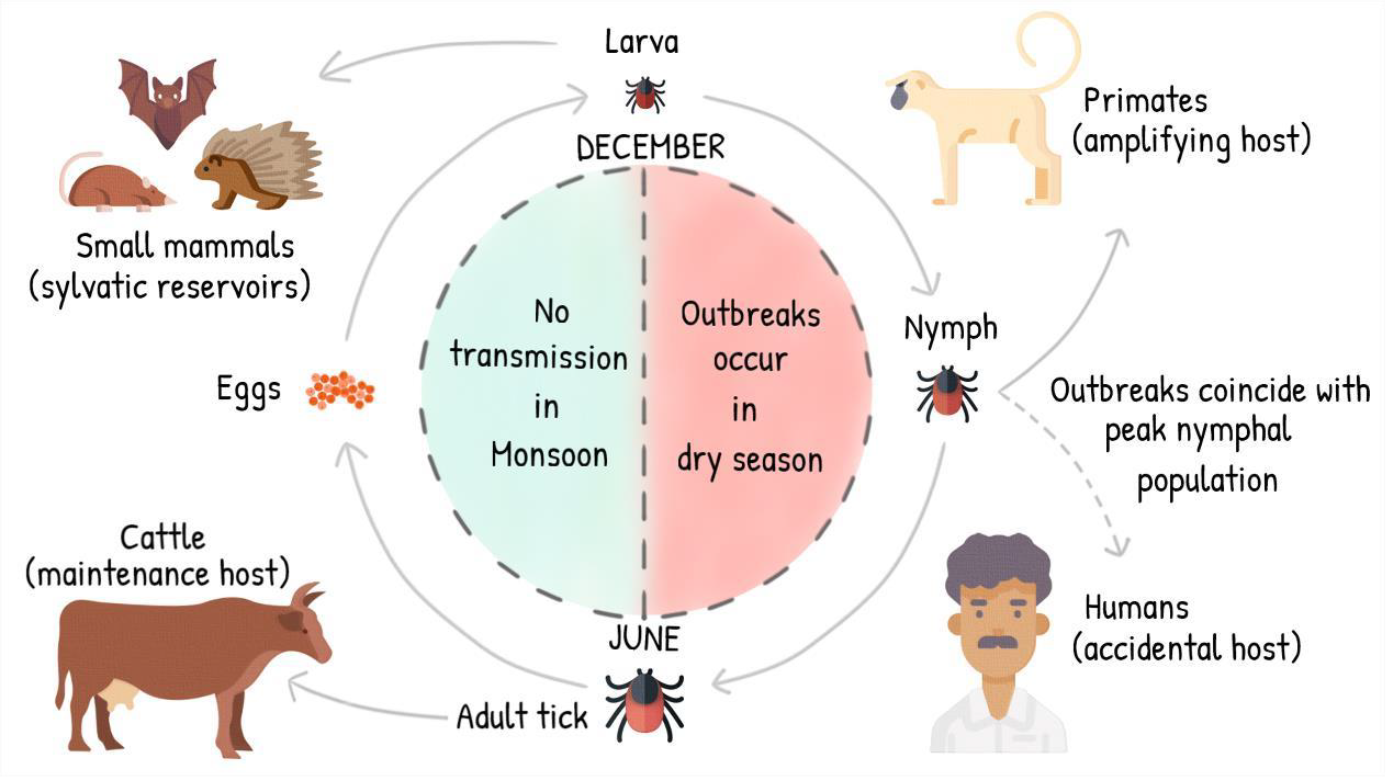
Transmission cycle of Kyasanur Forest Disease Virus in vector (*Haemaphysalis* ticks) and hosts (cattle, small mammals, primates and humans) with seasonal variations.

**Figure 2.**
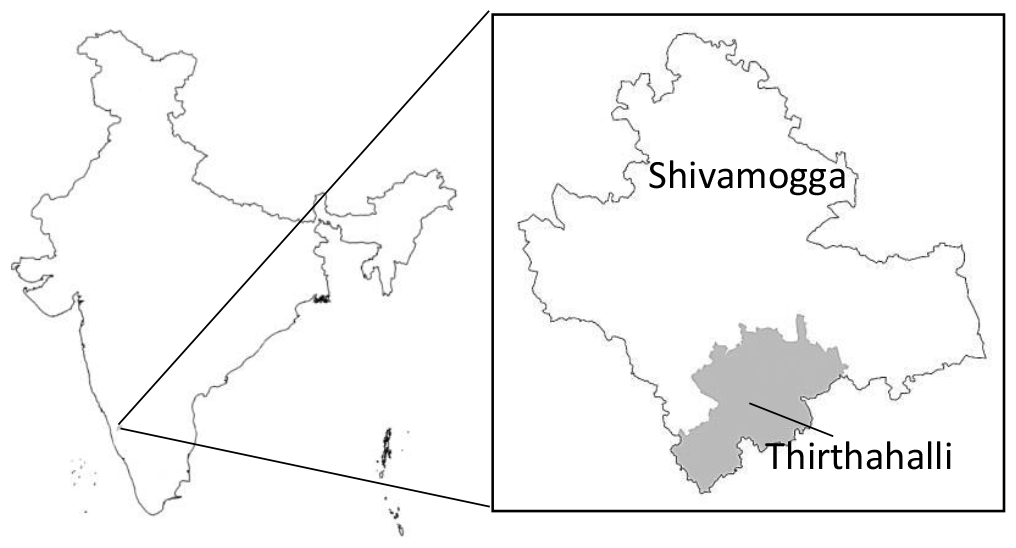
An overview map of Shivamogga district and Thirthahalli taluk (in inset), Karnataka state.

## Methods

We conducted a retrospective analysis of KFD surveillance data from November 2017 to May 2022. The operational definitions used by the KFD prevention and control program, Karnataka, are:

Suspected case: A person of any age presenting with acute onset of fever with any of the following: Headache/ Myalgia/ Prostration/ Generalized weakness/ Nausea/ Vomiting/ Diarrhoea/ Occasionally neurological/ haemorrhagic manifestations and associated with any of the following risk factors: Visit to endemic area* during past 2 weeks

Recent visit to unexplained monkey death area

(*Endemic area: An area which has reported human positive *and/or* monkey viscera positive *and/or* tick positive in the last 5 years).

### Confirmed case

A suspected case, which is laboratory-confirmed by any one of the following assays: l Detection of KFD Virus by real time RT-PCR or RT-PCR from serum or tissues.

l Detection of KFD Virus antibodies through anti-KFD IgM ELISA from serum.

KFD disease hotspot: a 50-feet radius around the area of an unusual monkey death Tick-pool: 20-30 ticks collected from one site

Sentinel site: Area which has reported human/ monkey/ tick positive in the last 5 years.

Random site: Area reporting fever cases in the present year/ high-tick density recorded during previous years.

Passive surveillance for human cases is ongoing throughout the year. Fever cases coming primary health centres, community health centres and private health facilities and exhibiting symptoms consistent with KFD are tested. Active surveillance is conducted during transmission season (November to June) around an arial radius of 5 KM surrounding the site where a human/ tick/ monkey positive for KFDV has been reported in the past five years. Surveillance workers conduct weekly house-to-house fever survey in the field from November to June, create a line list of suspected cases and send this list to the respective medical officer who arranges for sample collection. Government health facilities report human surveillance data on Integrated Health Information Platform (IHIP)/ Integrated Disease Surveillance Programme (IDSP) suspect case form (S-form). Blood samples are collected from suspected cases, packed and transported to VDL, Shivamogga. VDL tests the samples using human immunoglobulin enzyme-linked immunosorbent assay (IgM ELISA), reverse-transcription polymerase chain reaction (RT-PCR) or real-time RT-PCR. KFD vaccination is provided to eligible population residing around an arial radius of 5 KM surrounding the site where a human/ tick/ monkey positive for KFDV has been reported in the past five years.

District entomological team along with KFD field station staff collect tick pools from sentinel and random sites and test for presence of KFDV in VDL, Shivamogga, as per the operational guidelines^8^. Once a monkey death is reported, the respective gram panchayat team comprising a veterinary officer, forest guard, and health assistant visit the reported monkey death site, conduct necropsy, site decontamination, and send viscera samples to VDL Shivamogga or to NIV, Pune, to ascertain presence of KFDV. VDL compiles the data on human, tick and monkey positives from all districts and reports human positives in IHIP laboratory form (L-form).

Data was obtained from the Karnataka State Health Department and the district surveillance unit at VDL Shivamogga for the period January 2018 to December 2022. We obtained the annual cumulative list of lab-confirmed KFD cases for Shivamogga district and compared it with the annual KFD case burden in Thirthahalli taluk. We obtained the surveillance line-list of lab-confirmed (IgM or RT PCR) KFD cases for Thirthahalli taluk, Shivamogga district to calculate the annual and seasonal time-trend of cases. We obtained the surveillance data on monkey carcass site and tick pools, positive for KFDV, for Thirthahalli taluk to correlate with number of cases. We report Pearson’s correlation coefficient. We performed spatial analysis to identify clustering patterns and describe theprimary health centre (PHC)-wise disease burden. Epidemiological characteristics such as age, gender, place of disease onset, and outcome were analysed to identify high-risk groups.

## Results

From 2018 to 2022, Shivamogga district reported 609 cases and 17 deaths out of which 326 (54%) cases and six (35%) deaths were reported from Thirthahalli taluk (one of the seven taluks), with the proportion of cases from Thirthahalli out of all cases from Shivamogga district increasing from 44% in 2018 to 86% in 2022 (fig. 3).

**Figure 3.**
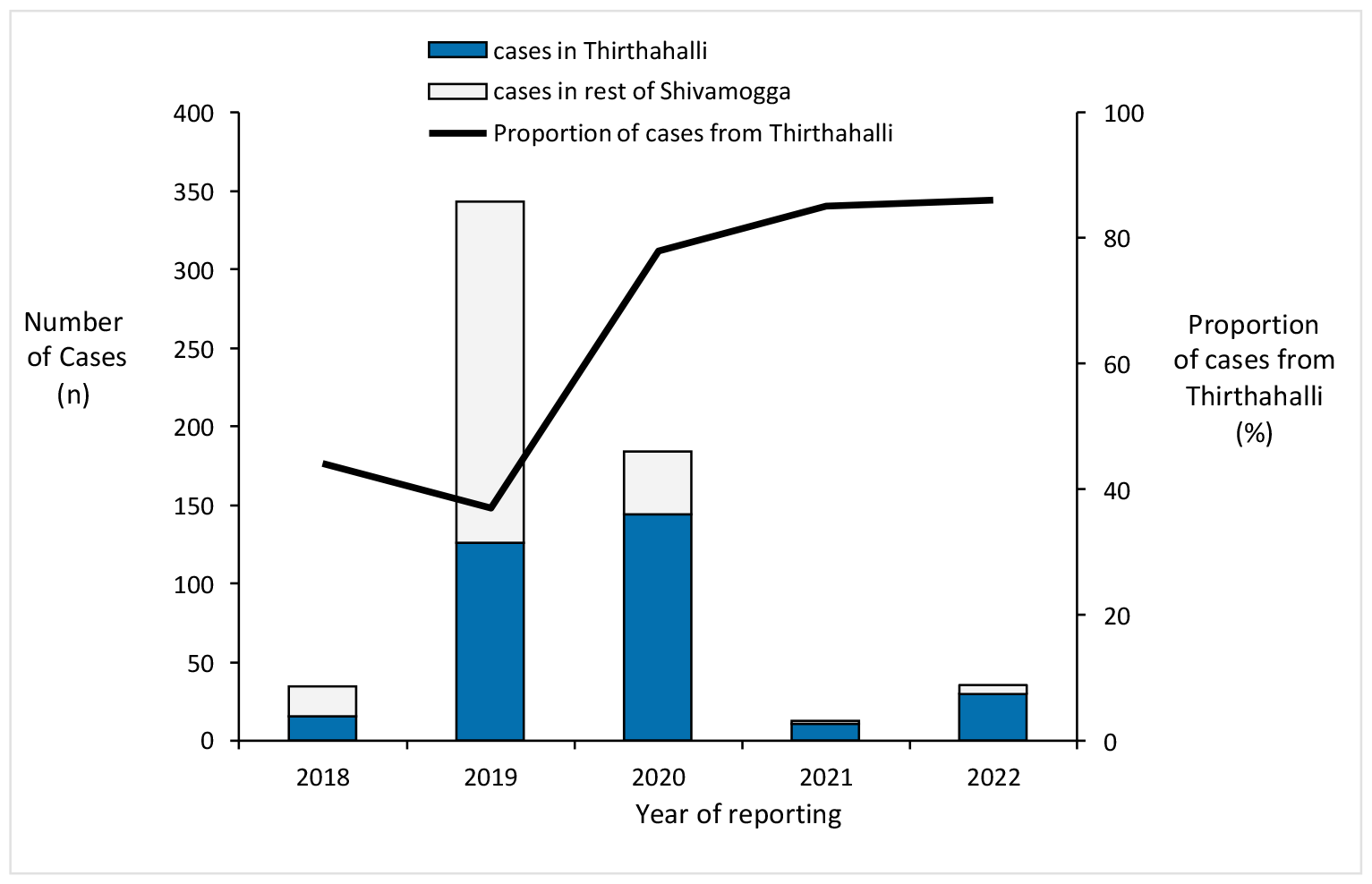
Trend of year wise lab-confirmed KFD cases in Thirthahalli taluk out of all KFD cases reported in Shivamogga district, Karnataka, 2018-2022.

During the study period, a total of 326 confirmed human cases of KFD were reported in Thirthahalli Taluk; all were lab-confirmed using either RT-PCR (93%) or IgM-ELISA (7%). The annual incidence rate of human KFD cases, calculated considering the population of Thirthahalli as 142,006 (Census 2011) was 45.9 per 100,000 population and varied between 11.3 per 100,000 population in year 2018 to 101.4 per 100,000 population in year 2020. The majority of cases were observed in males (188 cases, 58%), median age was 40 years (range 2-75 years) with an interquartile range (IQR) of 28 to 51 years. Cases occurred between the months of November and July, most cases occurring in the months of February to April (233 cases, 71.5%) with maximum cases reported in March (97 cases, 30%) as shown in figure 4. The case fatality rate was 1.8%.

**Figure 4.**
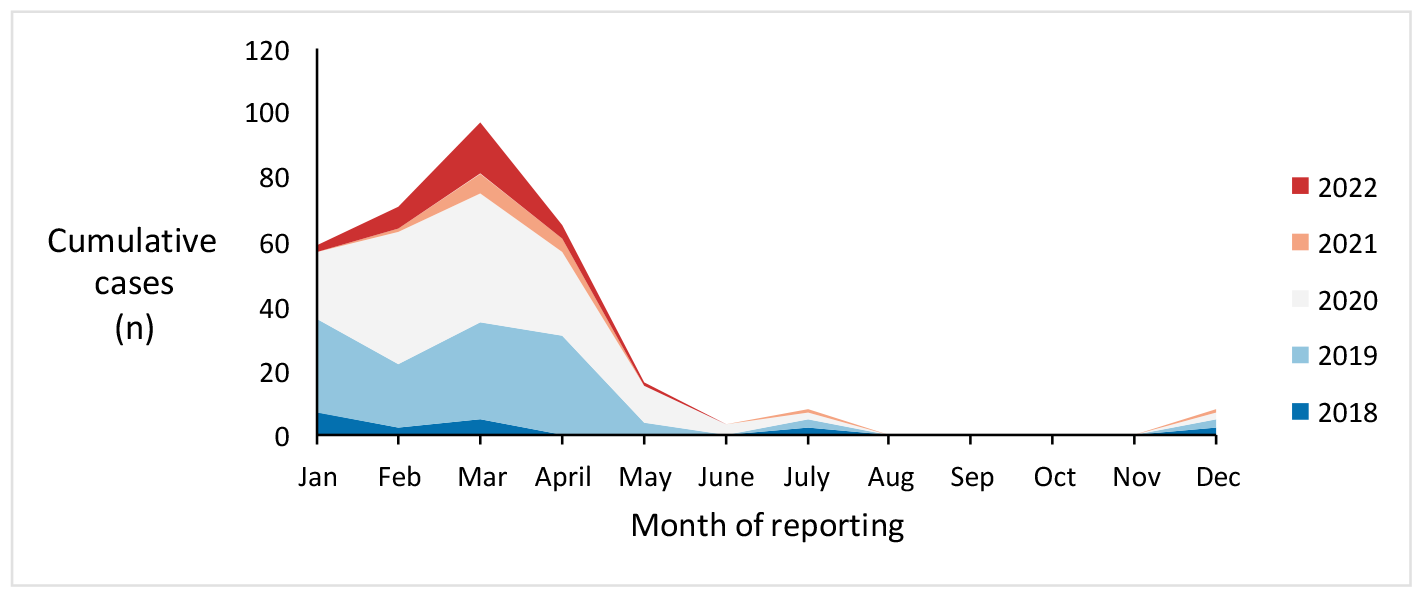
Seasonal trend of Kyasanur Forest Disease cases in Thirthahalli taluk, Shivamogga District, Karnataka, India, 2018-2022.

The maximum number of cases occurred in the years 2019 (n=127, (38%)) and 2020 (n=146, (44%)) crossing the outbreak threshold (20.3±8.7 calculated using the number of KFD cases reported from Thirthahalli in 2018, 2021 and 2022 as depicted in figure 5.

**Figure 5.**
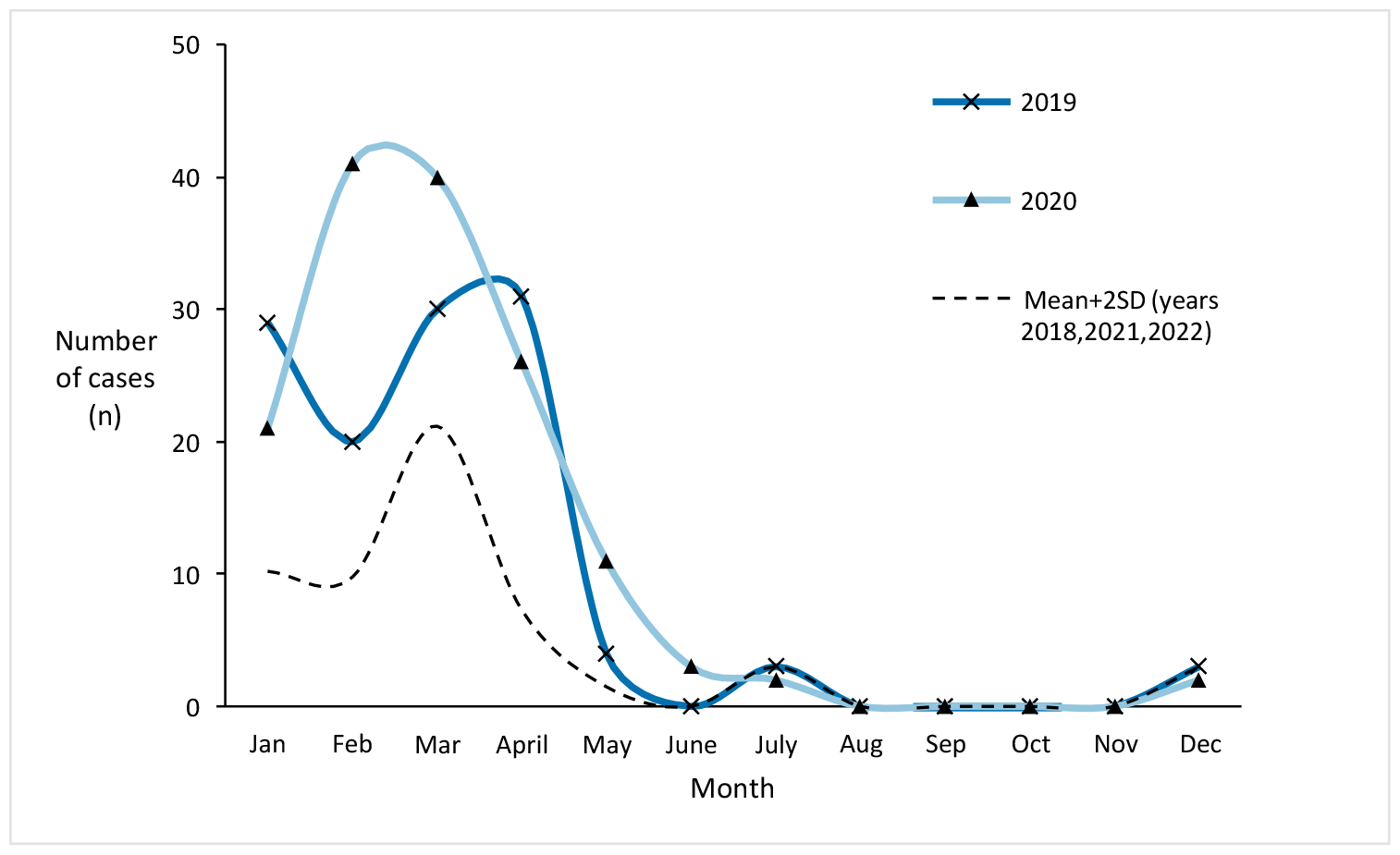
Consecutive KFD outbreaks in Thirthahalli taluk during the years 2019 and 2020.

The populations coming under six PHCs (Maluru, Kannangi, Mandagadde, Konanduru, Katagaru and Bettabasavani) contributed 84% of all cases reported from Thirthahalli from 2018 to 2022; and all deaths occurred in five of these PHCs. Four of these high-burden PHCs had tick-pools positive for KFDV but only PHC had monkey carcass positive for KFDV. One PHC had 3 positive tick pools, one positive monkey carcass but no human cases; 25% of the positive tick pools were reported from PHCs reported <1% cases. A correlation matrix (table 1) of PHC-wise distribution of human cases, human deaths, positive tick pools and positive monkey carcasses shows moderately strong correlation between human cases and positive tick pools (Pearson’s correlation coefficient, r=0.62) and between positive tick pools and positive monkey carcass (Pearson’s correlation coefficient, r=0.71); and weak correlation between human cases and positive monkey carcass (Pearson’s correlation coefficient, r=0.35).

**Table 1.** Primary Health Centre (PHC)-wise distribution of human KFD cases, human deaths andKFD hot-spots in Thirthahalli taluk, Shivamogga, Karnataka, 2018-2022.

| PHC | Human cases | Human deaths | Tick pools positive | Monkey carcasses positive |
| --- | --- | --- | --- | --- |
| **Maluru** | **83** | **1** | **0** | **0** |
| **Kannangi** | **56** | **1** | **19** | **0** |
| **Mandagadde** | **48** | **2** | **2** | **0** |
| **Konanduru** | **37** | **1** | **0** | **0** |
| **Katagaru** | **31** | **0** | **1** | **0** |
| **Bettabasavani** | **27** | **1** | **1** | **1** |
| Guddekoppa | 8 | 0 | 0 | 0 |
| Guthiyedehalli | 9 | 0 | 0 | 0 |
| JCH taluk hospital | 5 | 0 | 0 | 0 |
| Megaravalli | 3 | 0 | 0 | 0 |
| Yogimalali | 3 | 0 | 0 | 0 |
| Agumbe | 1 | 0 | 0 | 0 |
| Aralasurali | 1 | 0 | 2 | 1 |
| Devangi | 1 | 0 | 1 | 0 |
| Harogulige | 1 | 0 | 2 | 0 |
| Araga | 0 | 0 | 3 | 1 |
| Name not available | 12 | 0 | 0 | 0 |
| **Total** | **326** | **6** | **31** | **3** |

**Boldface** indicates high case-burden or death

During the study period, three monkey carcasses and 31 tick pools positive for KFDV were reported in Thirthahalli Taluk. Among these 94% tick pools were reported in the month of January and 6% in February; one monkey carcass was reported each in January, February and March in the years 2019 (n=2) and 2020(n=1).

The age group most affected by KFD were 45 to 64 years with the highest average annual incidence in the age group 45 to 49 years (8.4 per 100,000 population for 2018 to 2022) and 50-54 years (8.1 per 100,000 population for 2018 to 2022). The detailed age distribution of cumulative KFD cases and their average annual incidence in Thirthahalli taluk from 2018 to 2022 is shown in table 3. Among the six KFD case-patients who died, median age was 58 years (range 42-75 years), IQR 51 to 65 years and 83% were males.

**Table 2.**
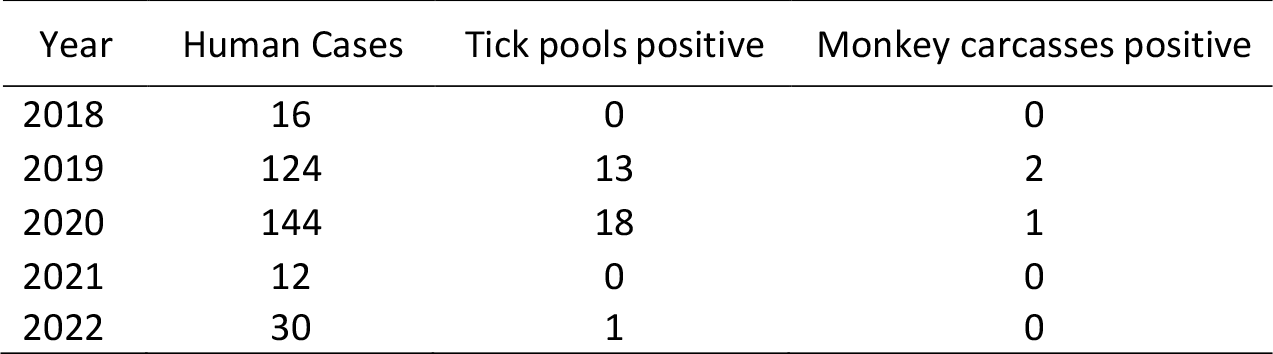
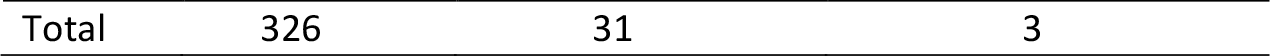
Year-wise comparison of human KFD cases, tick pools positive for KFDV and monkey carcasses positive for KFDV in Thirthahalli taluk, 2018-2022.

**Table 3.** Age-group wise annual average incidence of KFD cases in Thirthahalli taluk, Shivamogga, 2018-2022.

| Age group | Cumulative cases (2018-2022) | Population (Census 2011) | Average annual incidence (per lakh population for 2018-2022) |
| --- | --- | --- | --- |
| 0-4 | 9 | 129212 | 1.4 |
| 5-9 | 9 | 138103 | 1.3 |
| 10-14 | 11 | 162453 | 1.4 |
| 15-19 | 16 | 164421 | 1.9 |
| 20-24 | 21 | 164792 | 2.5 |
| 25-29 | 23 | 154169 | 3.0 |
| 30-34 | 30 | 130707 | 4.6 |
| 35-39 | 30 | 140421 | 4.3 |
| 40-44 | 29 | 123091 | 4.7 |
| **45-49** | **50** | **119078** | **8.4** |
| **50-54** | **36** | **89754** | **8.0** |
| **55-59** | **25** | **70402** | **7.1** |
| **60-64** | **17** | **58849** | **5.8** |
| 65-69 | 11 | 44556 | 4.9 |
| 70-74 | 5 | 23994 | 4.2 |
| 75-79 | 4 | 15527 | 5.2 |
| 80-84 | 0 | 10242 | 0.0 |
| 85-89 | 0 | 3694 | 0.0 |
| 90-94 | 0 | 1831 | 0.0 |
| 95-99 | 0 | 824 | 0.0 |
| 100+ | 0 | 469 | 0.0 |

**Boldface** indicates high average annual incidence

## Discussion

The results of our study indicate an ongoing endemic transmission of KFD in Thirthahalli, with the proportion of cases from Thirthahalli gradually increasing from 2018 to 2022, to contribute the majority of cases in Shivamogga district. Two KFD outbreaks occurred in consecutive years with the occurrence of cases following the established seasonal pattern. Around a quarter of the PHCs had high case burden among all PHCs in Thirthahalli taluk. All KFD deaths s occurred in PHCs with a high case burden and occurrence of human cases was moderately correlated with presence of tick pool positivity. Age groups most at risk for both the occurrence of the disease and mortality were the middle-aged adults.

Clustering of cases in specific areas of the taluk suggests localized transmission hotspots. Such areas may coincide with dense forest regions, highlighting the role of the forest ecosystem in the transmission dynamics of KFD. Similar to our study, Banerjee et al report high correlation between the number of human cases and infected tick pools over different years.^11^ However, Yadav et al argue that there is no correlation between human cases and positive tick pools based on their finding that positive tick pools were reported from areas reporting no human cases.^10^ Presence of KFDV in an area indicates the possibility of future outbreaks as monkeys and small mammals can transport the virus to surrounding areas. We recommend continued surveillance of tick pool positivity to map areas with potential for KFD spread in the future.

In our study, most of these tick hot spots were reported at least a month before the peak season of KFD and hence provided a window of opportunity of a few weeks to implement targeted community awareness, vector control measures and intensified surveillance in these areas. Climate variability, along with diversity of forest type and possession of indigenous cattle are known to be important factors influencing the distribution and life-cycle of ticks; and along with tick pool infectivity data, should be considered to map potential hot-spots and direct vector control measures.^12,13^

There is a significant fall in the number of cases from 2020 to 2021. This could be attributed to two factors. The reporting in 2021 could have been compromised due to COVID induced attrition in surveillance staff and hence many cases might have been missed due to gaps in surveillance or, this could be a genuine fall in cases as people might have not gone into forest due to COVID-induced restrictions on movement.

The predominance of cases in adult males of specific age groups in our study is similar to that reported in literature and may be associated with occupational or behavioural factors that increase exposure to forest or farming activities and subsequently increase exposure to tick bites.^14,15,16^ Emerging zoonotic diseases are known to disproportionately affect small holder farmers from marginalised communities whose residence and occupation is closely linked to forest.^17^ Asaaga et al argue that access to disease information improved the ability of these marginalised groups to use adaptive strategies and hence preventive interventions should take into effect the social determinants of health.^18^ We recommend targeted awareness campaigns and preventive interventions in these age groups engaging in occupation that puts them at increased risk of acquiring the disease, keeping in mind the social context of these populations.

## Conclusion

This study underscores the importance of continued tick surveillance and monkey carcass testing to map disease hot-spots for implementing targeted interventions. Effective control strategies should focus on raising community awareness and enhancing preventive measures for individuals at higher risk. As the KFD vector exists in the forest, it is vital to follow a one-health approach with collaborative efforts between health authorities, forest officials, entomologists, animal husbandry, public health researchers, and local communities for managing and containing the spread of KFD in the region.

## Data Availability

All data produced in the present work are contained in the manuscript

## Ethical statement

This study was conducted as part of a public health outbreak investigation to identify risk factors for Kyasanur Forest Disease, a public health problem, and to institute appropriate control measures as part of public health response by the Karnataka State Health Department and hence was deemed non-research activity and ethical approval was exempted. Secondary data was obtained from the Karnataka state health department after obtaining due permissions and analysis was performed after removing patient identifier details. We shall maintain this data securely for five years after completion of study. We shall not reveal the participant’s identity in any publication or scientific presentation.

## Acknowledgements

We would like to acknowledge the participation and support of the following people in carrying out this study.

Dr Srinivasa GN, Project Director, IDSP, Karnataka; Dr Sharath Chandra, State Veterinary Consultant, IDSP; Dr Darshan Narayan, Research Scientist, VDL, Shimoga; Dr Harshavardhan, Deputy Chief Medical Officer, VDL; Dr Rajesh Surgihalli, District Health Officer; Dr Mallappa, District Surveillance Officer; Dr Nataraj, Taluk Health Officer, Thirthahalli; Ms Bhuvaneshwari, Public Health Entomologist; Mr Annapachary, Multi-purpose health worker (MPW); Mr Lokesh,MPW and Mr Vinay, MPW.

